# The Accuracy of Price Transparency Files at Top-Rated US Hospitals

**DOI:** 10.1101/2024.11.19.24317568

**Authors:** Austin J. Triana, R. Leland Van Horn, Mason Alford-Holloway, Joseph A. Quintana, Arthur B. Laffer, R. Lawrence Van Horn

## Abstract

Recent U.S. policies require hospitals and health insurers to post payer-specific negotiated prices for services. Few studies, if any, have thoroughly investigated the accuracy of these payer-specific prices for individual hospitals. Our aim was to evaluate the concordance of prices using three datasets: 1) hospital price transparency, 2) payer price transparency data, and 3) Electronic Remittance Advice (ERA) claims data, reflecting actual payments. Evaluating transthoracic echocardiograms at 22 top-rated hospitals, we found high variability in prices and low concordance between datasets, as less than 30% of claim payments had a concordant price in the price transparency datasets. Deriving accurate price estimates for patients requires precise matching of the insurance plan, place of service, and fee type, all of which should be included in standardized reporting. This process highlights the complexity of insurance contracting, and it is not surprising that there is some disagreement in prices. Regardless, we are hopeful that highlighting variation and complexity in prices is a step towards a better system of healthcare payments in the United States.

## Introduction

Recent legislation aims to improve healthcare consumer information, including the Transparency in Coverage (TiC) Final Rule, No Surprises Act, and CMS Hospital Price Transparency Rule.^1–3^ These three policies aim to empower patients make more informed healthcare purchases. Most hospitals and payers now disclose negotiated rates in publicly available machine-readable files.^4,5^Although this price transparency data is now publicly accessible, the accuracy of the data remains uncertain, and significant challenges limit usability for consumers, providers, and researchers, including exceedingly large files and inconsistent formats.^6–9^

To date, there have been few attempts to validate this data, each with shortcomings in specificity to individual hospitals and payers.^10–12^ In this study, we Electronic Remittance Advice (ERA) claims data to evaluate the accuracy and concordance of price transparency data at 22 top-rated hospitals in the United States for a single service: transthoracic echocardiogram (TTE). We are uniquely positioned to validate these prices because ERA claims are the gold standard— they contain the exact paid amount for each transaction. In addition, the ERA data allowed us to consider other important pricing factors including site of service and claim type, to evaluate both professional and institutional fees. These factors gave us an unprecedented, granular view of pricing both within and across institutions.

## Data and Methods

### Sample & Data Sources

The sample included 22 hospitals in the US News & World Report Best Hospitals list.^13^ This diverse set of hospitals spans a variety of states and metropolitan service areas throughout the United States and has previously been studied in price transparency research.^7^

Hospital and payer price transparency data was obtained from the Turquoise Rate Sense dataset.^14,15^ Turquoise Health aggregates, compiles, and cleans machine readable files that are posted by hospitals and payers. Both the hospital and payer data were extracted from Turquoise Rate Sense in December 2023. The publicly available payer-specific negotiated rates posted by hospitals will be referred to as “hospital prices,” and rates published by insurance providers will be referred to as “payer prices.” We excluded rates that were given as a percentage of gross charge to avoid extrapolation.

The ERA claims data was obtained from Preverity, a healthcare data and analytics company. Preverity’s dataset includes roughly 60% of all Electronic Data Interchange (EDI) 837 commercial claims in the United States. Preverity combined both EDI and ERA data at the individual claim level to provide a complete description of the clinical care and payment including the date of transaction, common procedural terminology (CPT) code, quantity of services, location, provider, and payer, without patient-identified information. We analyzed commercial claims that occurred analyzed from January 1, 2022 to December 31, 2022 from the Preverity dataset for the top US hospitals. The paid amounts that are observed in claims data will be referred to as “claim payments.”

### Data Cleaning

For each hospital, a list of associated National Provider Identifiers (NPIs) was extracted from the American Hospital Directory and Definitive Healthcare and mapped to the 22 hospitals included in this analysis. Roughly 10% of billing NPIs were excluded because they were associated with multiple hospitals in the list (for example, Massachusetts General Hospital and Brigham and Women’s Hospital). In total, 105,998 unique billing NPIs were mapped to the 22 hospitals and were included in the analysis. Only claims with the associated CPT code 93306 were included.

A major challenge was to match payer names across the three datasets. Because there is no unique identifier, the payer name is often represented by a nonstandard string of text. Through an extensive data cleaning process, Turquoise created a map to identify payer class and payer type for the price transparency data.^16^ A similar process was then used for the ERA claims data to extract the payer using rule-based string extraction and manual labeling. Finally, the three datasets were merged on payer, hospital, and CPT code so that payer-specific rates could be compared. There were no inconsistencies in NPI numbers nor CPT codes. However, there were several inconsistencies in payer names. Any remaining payer names that could not be matched and merged were grouped into an “Unsorted” category. This methodology included a small degree of manual matching of insurance plans and payers. A different string extraction and labeling algorithms may lead to slightly different results, especially if not using the Turquoise Rate Sense network map.

In the ERA claims dataset, 11,787 claims for echocardiograms were found for the 22 hospitals in 2022. Each ERA claim was crossmatched with the corresponding EDI claim to extract other key data, including the place of service and claim type. Claims were sorted by payer and hospital. We did not capture any claims on two hospitals: UCLA and Barnes Jewish Hospital. We captured less than 10 claims on three hospitals: Massachusetts General Hospital, Vanderbilt, and Rush.

### Measuring Concordance

Concordance was evaluated by searching for matching payer-specific rates in each dataset. For each claim payment, we defined a “match” as a corresponding price in a price transparency file within $100 of the claim payment, a small margin to account for claim adjustments. Concordance was contingent on matching the hospital name and payer name. We did not include place of service in the concordance analysis. Ultimately, we were able to determine the percentage of claim payments that had a matching price in each of the price transparency files.

## Results

In the hospital and payer price transparency files, the listed price of an echocardiogram ranged from $1.3 to $34,424. In the ERA claims data, the median payment for the professional fee was $203 (Q1 = $116, Q3 = $245), and the median payment for the facility fee was $2682 (Q1 = 1829, Q3 = $4531). There was a ten-fold difference in median price between the cheapest and most expensive facility.

Figure 1 shows claim payments for echocardiograms for hospitals. Several prices were only observed once or twice, likely reflecting noise in the data due to a nominal adjustment for patient responsibility or a deductible. Thus, a sensitivity analysis was performed to filter out price points that were not observed on multiple occasions, removing 2616 claims.

**Figure 1:**
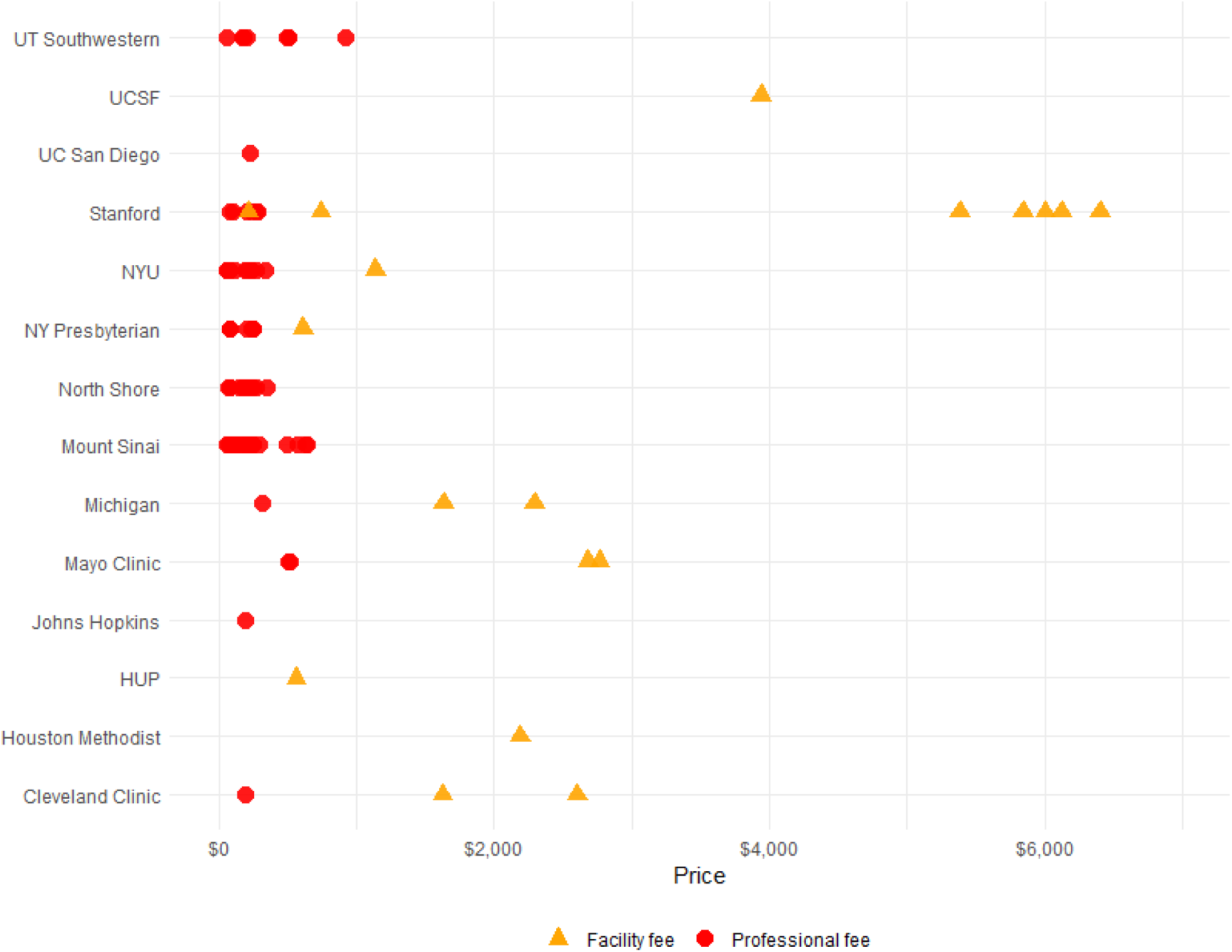
Prices of echocardiograms at top-ranked hospitals obtained from ERA claims data Claims data was obtained on 14 of the top-rated hospitals. Expanded hospital names are listed below. UCSF: University of California San Francisco Medical Center. NYU: New York University Langone Tisch Hospital. NY Presbyterian: New York Presbyterian Hospital Columbia and Cornell. Michigan: Michigan Medicine University Hospital. Mayo Clinic Hospital Rochester, Minnesota. HUP: Hospital of the University of Pennsylvania.

Figure 1 shows professional and facility fees for echocardiograms for 9,171 claims. These 9,171 claims fell on 69 distinct rates for professional fees and 18 distinct rates for facility fees. This graph demonstrates a few important points:

1. **Most services transact on a few distinct prices.** We found that 80% of claim payments fell on 87 distinct rates. For example, all the facility fee payments for Hospital of the University of Pennsylvania (HUP) and Houston Methodist fell on a single point in the ERA claims data. By comparison, we observed 643 unique rates in the hospital price transparency dataset and 727 unique rates in the payer price transparency set.
2. **Professional fees generally range from $100 to $300.** The median rate was $203 (Q1 = $116 and Q3 = $245). This pattern was generally seen across hospitals, and the variation is addressed later when discussing site of service.
3. **Facility fees are significantly higher than professional fees.** This corroborates prior analyses showing that facility fees tend to be much higher than professional fees.^17^
4. **There is large variation in facility fees.** Stanford and UCSF were the most expensive facilities by a large margin.

For each hospital, echocardiogram prices from all three data sources were graphed according to payer, as shown in Figure 2 and Figure 3. The hospital and payer prices (blue and green crosses) are shown as distinct prices with no relationship to volume. Figures 2 and 3 include low-volume claim payments to illustrate more detail than Figure 1.

**Figure 2:**
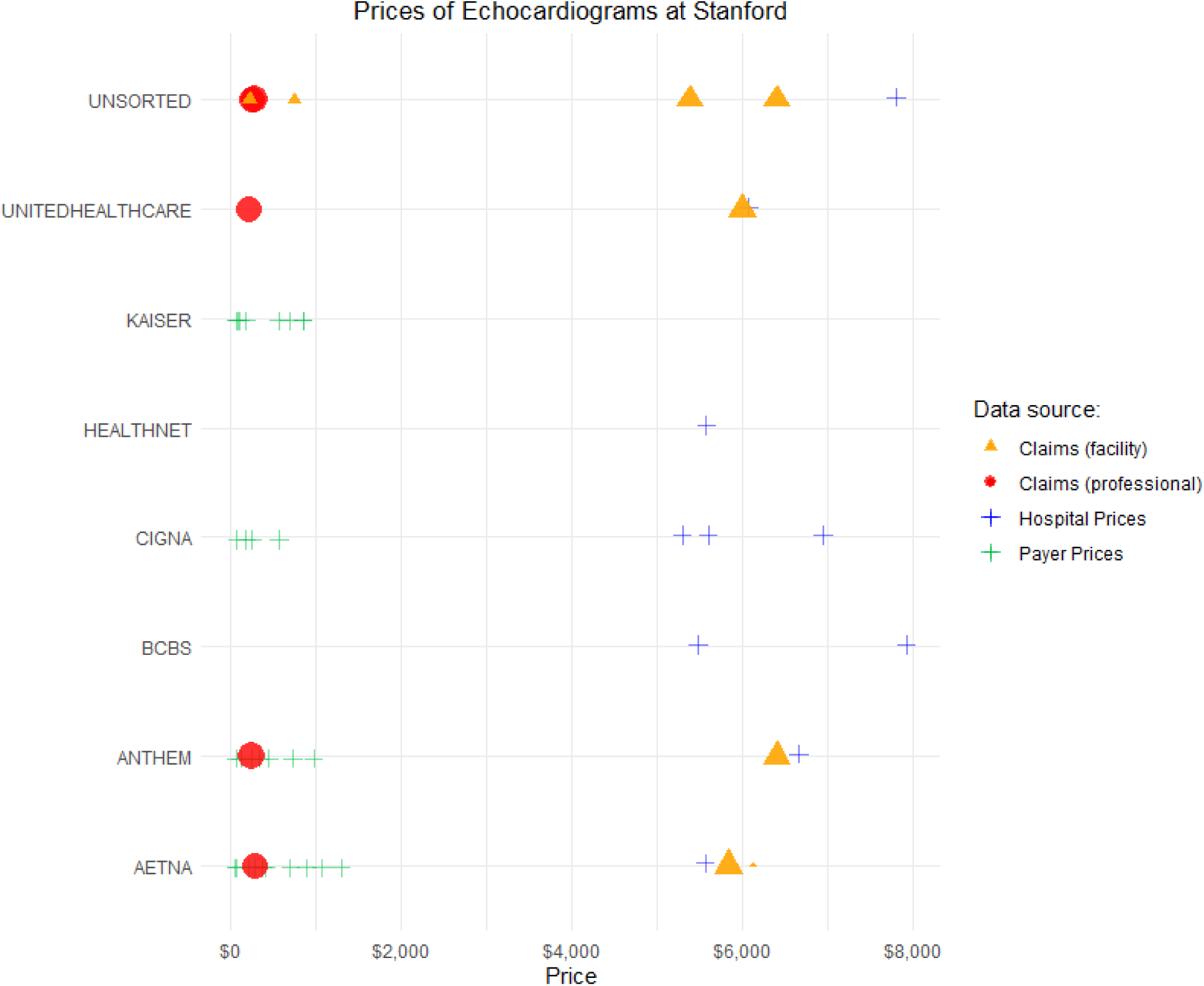
Payer-specific rates of echocardiograms at Stanford

**Figure 3:**
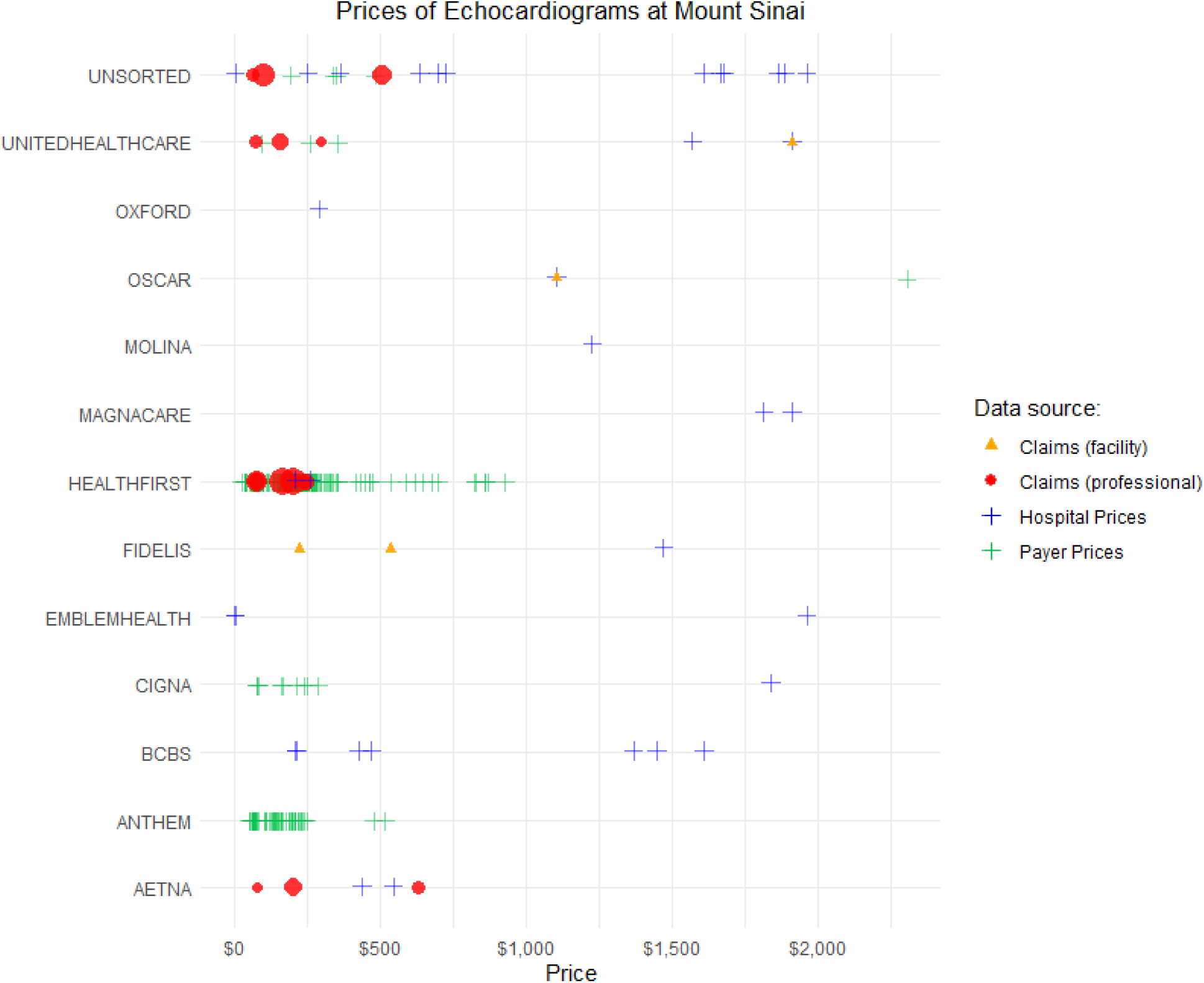
Payer-specific rates of echocardiograms at Mount Sinai

Figure 2 represents 1,228 claims observed in the ERA data, and Figure 3 represents 4,263 claims. These figures reveal that many publicly posted prices are not associated with any transaction volume. We suspect that there are many rates that may be contractually true but not practically relevant. For example, Hospital of the University of Pennsylvania (HUP) posted nine prices greater than $20,000 for an echocardiogram with a maximum of $34,424, none of which were observed in the ERA claim data. These may be similar to “dummy rates” or “ghost rates” and ultimately are not helpful for consumers.

In addition, the price transparency data appears incomplete. Looking at prices at Mount Sinai, we observe facility fees for United Healthcare at $1,800 and Oscar at $1,100, both of which are validated by a blue cross (hospital price transparency data). However, the two rates for Fidelis at $223 and $535 do not have any corresponding price, suggesting that the price transparency datasets may still be incomplete.

Figure 4 outlines measures of concordance between ERA claim payments and the price transparency datasets. For both hospital and payer datasets, less than 10% of claim payments had an exact matching price. Many of the claim payments observed were subject to small adjustments (most commonly patient responsibility or copay), so we widened the margin to search for “near matches” as above.

**Figure 4:**
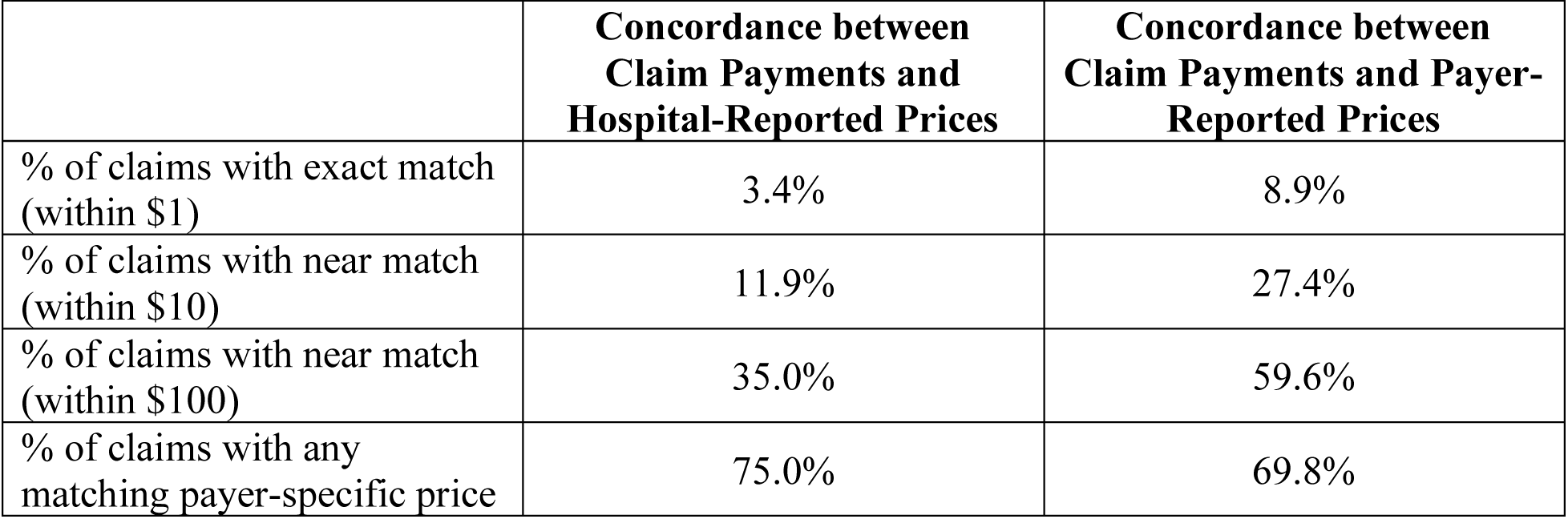
Sensitivity analysis of concordance measurement between ERA claim payments and price transparency datasets

## Discussion

Despite promising levels of concordance between the three datasets, we have shown that the current publicly available prices do not reflect the reality of the transacted payments. Decades of insurance contracting has resulted in a convoluted, byzantine structure in which it is highly challenging to derive a simple price that is communicable to a layperson (or nurses and physicians). The goal of healthcare consumerism is admirable, but the current system makes it extraordinarily challenging for anyone to extract actionable price information.

The purpose of this work was to investigate concordance of publicly available prices, and despite several challenges, we have shown that it is possible elicit and validate the true prices of services. Prior work has described wide price variation of payer-specific rates.^9^ On the other hand, we have shown that echocardiograms often transact on a distinct price once factoring the hospital, payer, plan, site of service, and type of payment (professional or facility fee). We found that less than 50% of claim payments had a matching price within $10 in the payer and hospital transparency datasets. We suspect this low concordance is in part due to claim adjustments but also reflects the intricate structure of medical billing and inconsistent data formats. It appears that payers have been more comprehensive; however, there are still major gaps in both data sets.

Furthermore, this work confirms significant inter-facility price variation. As shown above, payer-specific prices at a single a hospital cluster at distinct prices. However, prices between two hospitals may be wildly different. Perhaps the most drastic example is echocardiograms at Stanford. United Healthcare’s facility fee of an echocardiogram at Stanford is over three times higher than United Healthcare’s facility fee at Mount Sinai, suggesting it may be economical for a patient to fly from San Jose to New York City (possibly first class) for a simple diagnostic test.

These findings also have major policy implications:

1. Price transparency machine readable files need to be standardized. The complexity and variation of the files posted by payers and hospitals has proven to be exceedingly difficult for experts to handle, and practically impossible for consumers to parse. No files should report price as a percentage of the gross charge.
2. Price transparency files need to include site of service and claim type. This detail was present in the payer dataset but not in the hospital dataset. It is critical to distinguish facility fees from professional fees, and we have demonstrated that prices vary depending on the site of service.
3. “Ghost rates” and outlier prices should be investigated, and it may be reasonable to enforce fines for inaccurate or imprecise data. Our analysis suggests that the price transparency data includes prices that are never observed in real transactions, which is misleading for patients.

There are some limitations to this current study. This sample of hospitals and claims is neither representative nor comprehensive, limiting generalizability to other healthcare services and hospitals. In addition, the ERA claims data was imperfectly labeled. Between 30-60% of claims do not include proper labeling regarding place of service and patient obligation modifiers, further contributing to data loss. It is possible that similar noise exists in the price transparency machine readable files; however, the cleaning and handling of these files was performed by a third party and not these authors. Finally, it is possible that some of the contractual agreements changed in a year from 2022 to 2023, when the price transparency files were extracted from Turquoise.

## Conclusion

This work is an unparalleled view into real transaction prices for healthcare services. We have demonstrated the complexity of claim payments and highlighted major challenges to price transparency in healthcare. Wide price variation in reported prices is problematic, as this does not reflect the true distribution of claim payments. Instead, this study has demonstrated that claim payments transact on a few distinct payer-specific rates at each hospital.

While there are major limitations to both the current hospital and payer price transparency datasets, the healthcare industry has made rapid progress toward improving the consumer experience. We believe that price transparency is a process—a step towards a better system of healthcare payments in the United States, one that empowers patients. To disentangle decades of complexity in medical claims for the consumer’s understanding is not only a noble task; it is a necessity.

## Data Availability

All data produced in the present study are available upon reasonable request to the authors.

## Funding

None

## Disclosures

R. Leland Van Horn is an employee of Preverity.

R. Lawrence Van Horn is founder and CEO of Preverity. In addition, he has been Governance Committee Chair and Compensation Committee Member for Community Health Care Realty Trust (NYSE: CHCT). He is Board Chair for Savida Health and Advanced Behavioral Solutions. He is a member of the CEO Council for Council Capital and advisory boards for Harpeth Capital, Sidecar Health, and Abridge Health, and he is an operating partner with Whistler Capital Partners, LLC.

## Notes

### Funding Statement

This study did not receive any funding

